# Food Security after Nuclear Winter: A Preliminary Agricultural Sector Analysis for Aotearoa New Zealand

**DOI:** 10.1101/2022.05.13.22275065

**Authors:** Nick Wilson, Marnie Prickett, Matt Boyd

**Affiliations:** University of Otago, Wellington, New Zealand; Adapt Research Ltd, Reefton, New Zealand

**Author notes:** Corresponding author: Prof Nick Wilson.

## Abstract

**Aim:** We aimed to estimate the current dietary energy content of food exports for Aotearoa New Zealand (NZ) and food security after “nuclear winter” scenarios following a nuclear war.

**Methods:** From published sources we estimated dietary energy available from the major domains of food exports, with adjustments for wastage. The impacts on food production in NZ after three nuclear winter scenarios were based on those published in *Nature Food* in 2022 and from an earlier NZ Planning Council study.

**Results:** Current major food exports are equivalent to 3.9 times current dietary energy intakes for all NZ citizens ie, 34,100 kJ (8150 kcal) per person per day. Exported dairy products were estimated to be able to provide 338% of this energy intake, followed by exports of: meat (34%); fruit (8.6%), alcohol (4.8%), marine products (4.6%), and vegetables (2.7%). After the various nuclear winter scenarios considered (minimal to severe), food production available from diverted exported foods was estimated to still be 3.6 to 1.5 times current daily energy intakes.

**Conclusions:** This analysis suggests that NZ could have excess food production capacity, even after a severe nuclear winter scenario. But substantial further research is needed to clarify agricultural impacts and the role of nuclear war impacts on the interlinked domains of energy, transport, manufacturing, finance, industrial materials, trade and societal functioning.

## Introduction

The survival of human civilization, or its continued flourishing, could be threatened by a global catastrophe abruptly reducing sunlight reaching the earth.^12^ Such scenarios could include nuclear winter arising from a nuclear war;^3^ large volcanic eruptions^45^; and a large asteroid/comet impact.^6^ Resulting global climate impacts could include a drop in mean temperature that would limit food production possibly causing a catastrophic global food shock.^1^ Modelling studies indicate that the impacts of such catastrophes are likely to be highly heterogeneous around the world.^37^ It is sometimes thought, for example, that islands in the Southern Hemisphere might do better than countries in Northern Hemisphere landmasses during nuclear winter.^8,9^

Collectively these threats are not improbable. Estimates for the annual probability of inadvertent nuclear war include 1% ^10^, or in the 0.3% to 3% range.^11^ However, the uncertainty may even be higher and could be increasing with the Russian invasion of Ukraine in 2022. Another consideration is the risk of major volcanic eruptions on the scale of the Indonesian volcano Tambora in 1815. This eruption cooled global climate and contributed to famines in parts of Europe, India and China.^12^ Eruptions of a scale of Tambora or greater occur around 1.6 times per millennium,^13^ equivalent to around a one in six chance per century.^5^ But some estimates put the overall likelihood of a catastrophic global food shock (of >10% loss of production) at 80% this century.^14^

Other catastrophic scenarios include severe pandemics (such as those arising from engineered bioweapons), or global industry-disabling scenarios such as coronal mass ejections or electromagnetic pulses,^15^ that might collapse the global economy and isolate remote nations like Aotearoa New Zealand (NZ).

Vastly greater efforts are needed to prevent and mitigate all these threats to humanity, but it is also prudent to consider what happens if prevention fails for nations and the international community to plan effectively. In terms of catastrophic pandemics, past work has suggested that isolated island refuges could have a role in ensuring human survival. NZ appears relatively well placed in terms of survivability compared to other island nations, with it ranked first in one study^16^ and only behind Australia in others.^17,18^ Self-sufficiency on dimensions including food supply would have to be assured, although the previous work also reported the large per person food production capacity of NZ.^17,18^

Past work for NZ on the impact of nuclear war is substantively out of date as it was done in the 1980s eg, by the Commission for the Future,^19^ the NZ Planning Council^20–24^ and others.^25^ Since this time NZ society has changed in many ways, including the expansion of agricultural production, the growth in food exports, population growth, and technological developments. The scientific understanding of the impacts of abrupt sunlight reduction catastrophes has evolved as well.^26–29^

Given the above, we aimed to begin to explore the food security issues for NZ after such catastrophes by describing the current food export economy as well as the potential food supply when accounting for the climate impacts of a nuclear winter. In this preliminary work, we focused on the food *export* sector as the data appeared to provide a much clearer picture of food production than the far more diverse and complex domestic food production sector.

## Methods

### Business-as-usual dietary energy intake of New Zealanders

Adult,^30^ and child,^31^ nutrition survey data for NZ was used to estimate dietary energy intakes for these population groups by sex. These estimates were then multiplied by the relevant estimated NZ population sizes for the fourth quarter of 2021^32^ to estimate the national dietary energy intake.

### Food export analyses

Food export weights were obtained from the NZ Harmonised System for export data^33–35^ and five-year annual averages were calculated for the five-year period ending June 2020. We ignored food export categories that were under a 10,000 tonnes annual average in volume.

### Food wastage adjustments

The adjustments for potential food wastage are shown in Table These used data from the United States, United Kingdom and Europe. Although work on waste has been conducted in NZ,^36^ this work has not identified waste as a proportion of specific food products taken into the household.

### Dietary energy data

For the food exports that were available for intake at the household level after unavoidable wastage (inedible components) and avoidable wastage, we calculated the food energy available. This was done by matching typical foods within each food export category using the NZ Food Composition Database.^42^ We used representative foods within each category eg, in the cheese category: “cheddar cheese, code F1015”; and for the apple category: “Royal Gala, code L1150”. A full list of codes and the Excel spreadsheet is available on request to the authors.

### Nuclear winter scenarios

There is much uncertainty concerning the global impacts of nuclear war and if “nuclear winter” impacts would even occur. Nevertheless, to inform potential planning purposes we considered the three scenarios outlined in Table 2. The key driver of climatic impacts in these scenarios is stratospheric soot injection following nuclear weapon explosions on targets in the Northern Hemisphere. This stratospheric soot results in global cooling which impedes food crop production. In all these scenarios we assumed an effective end to NZ’s food trade with other countries (including Australia) for both exports and imports. This was also the approach taken in previous NZ research.^43^

**Table 1.** Assumptions around inedible fractions of foods and typical avoidable food wastage (for food previously prepared for export which was then assumed to be diverted to the domestic market after a global catastrophe)

| Major food export group (if over 10,000 tonnes per year) | Estimated food wastage during transport from food production facility to retailers and then to households [A] <sup>a</sup> | Estimated inedible fraction of exported food weights (unavoidable waste) [B] | Estimated avoidable wastage for foods entering the household [C] |
| --- | --- | --- | --- |
| Dairy products | 1% | 0% | 8% (WRAP) |
| Marine food products | 1% | 0% (most exports assumed to be filleted) | 8.5% (mid-point of 2 estimates: 6% (NRDC) and 11% (WRAP)) |
| Alcohol | 1% | 0% | 8% (value for all types of drinks) (WRAP) |
| **Red meat products** |  |  |  |
| Beef and veal | 1% | 18% (bone) <sup>b</sup> | 8.5% (mid-point of 6% (NRDC) and 11% (WRAP)) |
| Lamb and mutton | 1% | 16% (bone) <sup>c</sup> |  |
| **Fruit** |  |  |  |
| Apples | 5% | 12% <sup>d</sup> | 14% (mid-point of 10%<sup>37</sup> and 18% (WRAP) for fruit as a grouping) |
| Avocados | 8% | 26% <sup>d</sup> |  |
| Kiwifruit | 6% | 18% <sup>d</sup> |  |
| **Fresh vegetables** |  |  |  |
| Onions (dry) | 6% | 11% <sup>d</sup> | 16.5% (mid-point of 13%<sup>37</sup> and 20% (WRAP) for fresh vegetables as a grouping) |
| Potatoes (actually a mix of fresh & frozen) | 7% | 16% <sup>d</sup> |  |
| **Processed vegetables** |  |  |  |
| Peas (frozen) | 1% | 0% | 14% for processed vegetables (WRAP) |
| Sweetcorn (frozen) | 1% | 0% |  |

**Notes:**
<sup>a</sup> Retail waste was estimated from wastage in a US study by the National Resources Defence Council (NRDC)<sup>38</sup> (Table 62 in this NRDC publication). This indicated that the weight of food waste at the levels of “food wholesalers and distributors” and “grocers and markets” was 20% that of the weight of waste at the residential level (we combined the amounts for the three cities in the study). We applied this value to the fresh fruit and vegetable categories (ie, multiplying the 20% with the summed values in columns [B] and [C]). For processed foods we could not identify specific values but assumed it would be much less than for fresh produce. So we used a guesstimate of 1% to account for some small proportional loss due to expired “use-by-dates”, accidental damage and refrigeration failures.
<sup>b</sup> We ignored boneless meat exports and just used a value for the proportion of bone in a study of NZ beef (hind quarter, “commercial composition”) at 18% (Table 2 in Bass et al<sup>39</sup>). Of note is that food can be extracted from bones (eg, to make soup) so that in disaster circumstances these are a potential food source.
<sup>c</sup> We ignored boneless meat exports and just used a value for lamb carcasses of 16%.<sup>40</sup>
<sup>d</sup> Inedible fractions from an average of three European studies as per De Laurentiis et al.<sup>37</sup> Avoidable food waste in the household for the European Union was 5 kg/person/year out of 52 kg of fruit taken into the household (10%). For vegetables the equivalent value was 13% (9/71). Some of these values are probably conservative eg, the 16% value is for fresh potatoes, and would be lower for the processed frozen potatoes that are a component of NZ’s exports.
NRDC: National Resources Defence Council, US wastage data.<sup>38</sup>
WRAP: Data for the UK (Table 51 in this Report<sup>41</sup>).

**Table 2.**
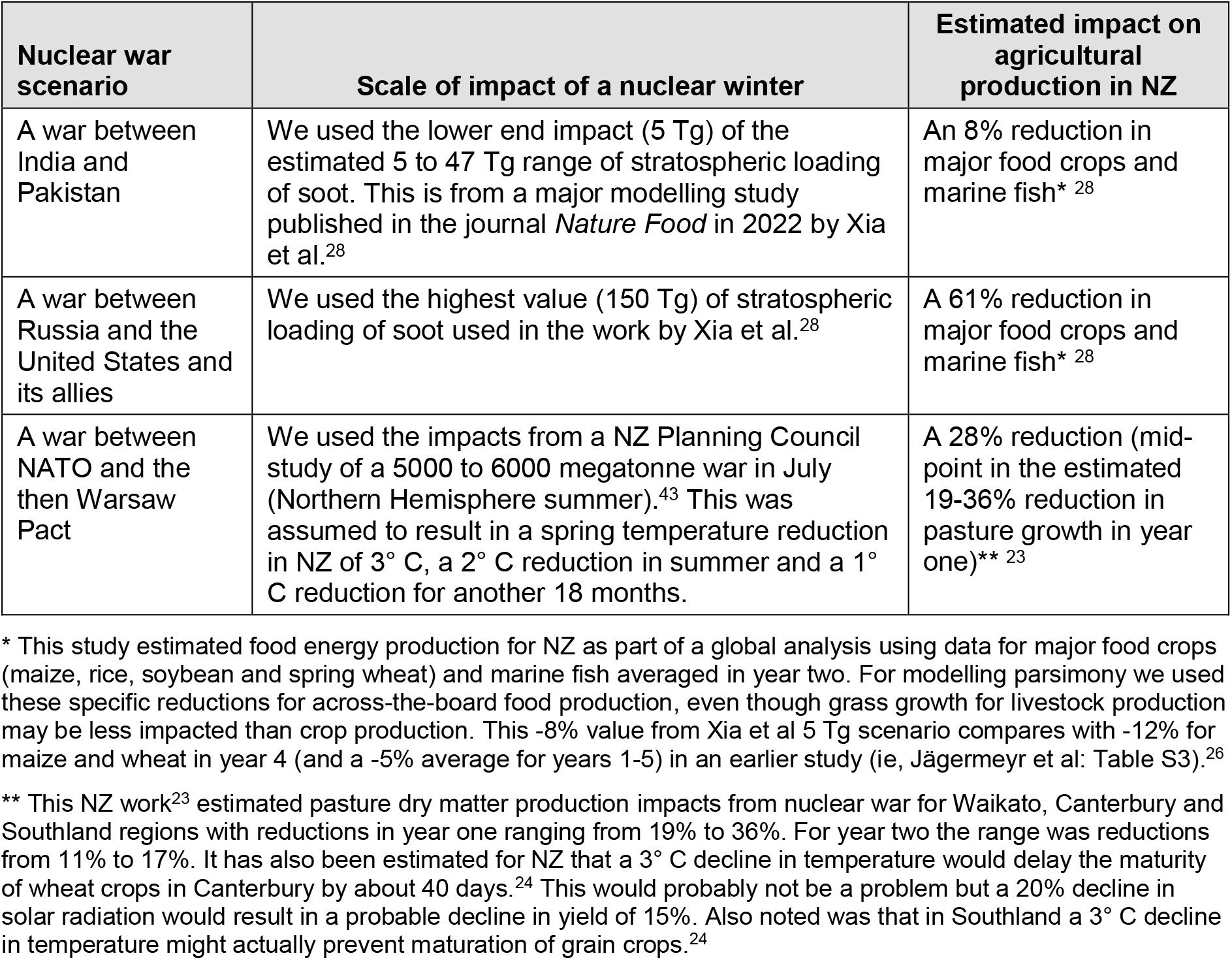
Three nuclear war and nuclear winter scenarios considered in this study

### Results

### Business-as-usual dietary energy intake of the NZ population

The estimated dietary energy intake of the entire NZ population was estimated at 44.4 billion kJ per day (8686 kJ per person per day; Table 3).

**Table 3.**
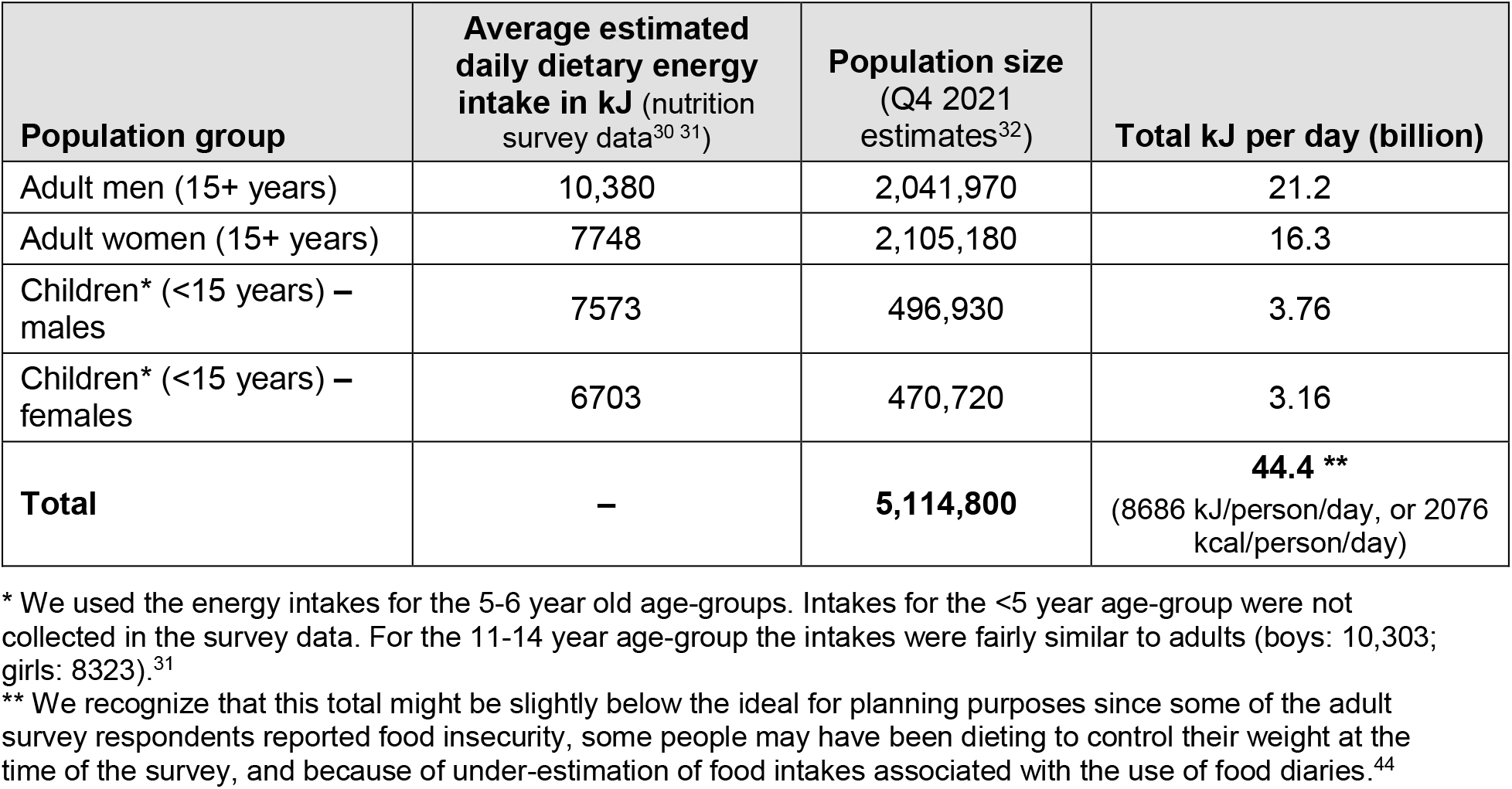
Estimated daily dietary energy intake of the total NZ population

^*^We used the energy intakes for the 5-6 year old age-groups. Intakes for the <5 year age-group were not collected in the survey data. For the 11-14 year age-group the intakes were fairly similar to adults (boys: 10,303; girls: 8323).^31^** We recognize that this total might be slightly below the ideal for planning purposes since some of the adult survey respondents reported food insecurity, some people may have been dieting to control their weight at the time of the survey, and because of under-estimation of food intakes associated with the use of food diaries.^44^

### Food energy availability from diverted exports

The estimates for the major food exports are detailed in Table 4 and summarised in Table 5. After food wastage adjustments, dairy products were estimated to provide 338% of all the current dietary energy, followed by: meat (34%); fruit (8.6%), alcohol (4.8%), marine products (4.6%), and vegetables (2.7%). Overall, these exports were estimated to provide 3.9 times dietary energy intakes for the whole NZ population (or 34,098 kJ [8150 kcal] per person per day).

**Table 4.** Food exports and available dietary energy estimates for NZ after adjusting for unavoidable food waste (inedible components) and avoidable food wastage (as per Table 1)

| Food export category (if over 10,000 tonnes per year) | Average annual exports (tonnes) – 5-year average ending June 2020* | Greatest contributor to weight | Daily food energy equivalents (kJ/day)* [% of daily required for all NZ population, adjusted for waste] | Further details |
| --- | --- | --- | --- | --- |
| **Dairy products (ordered by potential % of total dietary energy)** |  |  |  |  |
| Total milk, powder | 1,895,691 | Code=0402210019; “Dairy produce; whole milk powder, concentrated, not containing added sugar or other sweetening matter, of a fat content exceeding 1.5% (by weight), n.e.c. in item no. 0402.21” | 218% | Harmonised systems “dairy products” AND “milk”. Exports recorded in kgs. |
| Butter | 470,001 | Code=0405100001; Dairy produce; derived from milk, butter, unsalted | 82% | Harmonised systems “dairy products” AND “butter” |
| Cheese | 336,226 | Code=0406900011; Dairy produce; cheese, cheddar (other than in tins, not grated, powdered or processed) | 33% | Harmonised systems “dairy products” AND “cheese” |
| Total milk, liquid | 222,131** | Code=0401200901; Dairy produce; milk and cream, not concentrated, not containing added sugar or other sweetening matter, of a fat content, by weight, exceeding 1% but not exceeding 6%, UHT milk | 3.2% | Harmonised systems “dairy products” AND “milk”. Exports recorded as litres. |
| Other dairy product, including yoghurt, buttermilk, whey | 140,069 | Code=0403901901; Dairy produce; buttermilk powder, produced by a spray process, concentrated or sweetened, with or without flavouring, added fruit, nuts or cocoa | 1.3% | Harmonised systems “dairy products” remaining not fats |
| **Meat products** |  |  |  |  |
| Lamb and Mutton, including edible offal and processed meats | 398,267 | Code=0204420001; Meat; of sheep, lamb cuts with bone in, frozen (excluding carcasses and half-carcasses) | 18% | Harmonised systems “meat” AND “bovine” and “offal” AND “sheep” but not fats |
| Beef and veal, including edible offal and processed meats | 473,405 | Code=0202300001; Meat; of bovine animals, beef cuts according to the NZ Meat Producers' Board definition, of cow, steer and heifer, boneless, frozen | 16% | Harmonised systems “meat” AND “bovine” and “offal” AND “bovine” but not fats |
| **Marine food products** |  |  |  |  |
| Fish | 185,825 | Hoki | 3.3% | Exports Summary data – Quantity of Principal Exports |
| Molluscs | 61,106 | Code=0307430013; Molluscs; squid, frozen, whole | 1.3% | Harmonised systems “molluscs” |
| **Fruit** |  |  |  |  |
| Kiwifruit | 512,435 | 0810500019; Fruit, edible; kiwifruit, green fleshed, fresh | 4.5% | Harmonised systems “kiwifruit” |
| Apples | 362,537 | Code=0808100042; Fruit, edible; apples, Royal Gala, fresh, whole fruit | 3.4% | Harmonised systems “apple” |
| Avocados | 18,508 |  | 0.7% | Harmonised systems “avocado” |
| **Vegetables** |  |  |  |  |
| Onions | 175,110 |  | 1.0% | Harmonised systems “onions” |
| Potatoes (fresh, frozen or otherwise prepared) | 97,334 |  | 0.9% | Harmonised systems “potatoes” |
| Peas (frozen) | 38,507 |  | 0.7% | Harmonised systems “vegetable” search |
| Sweetcorn (frozen) | 11,270 |  | 0.2% | Harmonised systems “vegetable” search |
| **Other (converted from litres to tonnes)\*\*** |  |  |  |  |
| Wine | 257,006 |  | 4.4% | Harmonised systems “wine” |
| Beer | 33,307 |  | 0.2% | Harmonised systems “beer” |
| Spirits*** | 12,104 |  | 0.2% | Harmonised systems “spirits” |
| **Totals** |  |  |  |  |
| Total kJ per NZ citizen | – |  | 34,098 per NZ citizen per day | As per population groups in Table 1 |
| Excess factor relative to current intakes | – |  | 3.93 times dietary energy intakes | See Table 1 |

**Notes:**
\* We used the five-year average for exports between July 2016 to June 2020 to avoid too great an impact from Covid-19 disruptions.
\*\* The export values were converted from litres to tonnes using the following values: Assumed density of: liquid milk products at 1.035 kg/L; wine at 1.011 kg/L; spirits at 0.989 kg/L; and beer at 1.030 kg/L.
\*\*\*Export volumes recoded in “L.Alc” converted to litres using the lowest percentage alcohol in the category range.

**Table 5.** Daily dietary energy provided by major food export categories relative to the daily dietary energy intakes of the current NZ population and after three nuclear winter scenarios

| Major food export group (from Table 4) | Weight of annual food exports in tonnes (from Table 4) | Percentage of total NZ population intake (business-as-usual) (%) | Percentage of total NZ population dietary energy intake from diverted food exports after various nuclear winter scenarios (as per Table 2) (%) |  |  |
| --- | --- | --- | --- | --- | --- |
|  |  |  | 8% reduction in agriculture | 28% reduction in agriculture | 61% reduction in agriculture |
| Dairy products | 3,064,118 | 338% | 311% | 243% | 132% |
| Meat products | 871,672 | 34.1% | 31% | 25% | 13.3% |
| Fruit | 893,480 | 8.6% | 4% | 3% | 3.3% |
| Alcohol | 302,417 | 4.8% | 8% | 6% | 1.9% |
| Marine food products | 246,931 | 4.6% | 3% | 2% | 1.8% |
| Vegetables | 322,221 | 2.7% | 4% | 3% | 1.1% |
| **Total** | **5,700,839** | **393%** | **361%** | **283%** | **153%** |
| **Total kJ available** | – | 63,658,104 | 58,565,456 | 45,833,835 | 24,826,661 |
| **Total kJ per person per day available** | – | 34,098 | 31,370 (7498 kcal) | 24,551 (5868 kcal) | 13,298 (3178 kcal) |

### Impact of nuclear winter scenarios

The impacts on food availability from currently exported food after the various nuclear winter scenarios (as detailed in Table 2), are shown in Table 5. In the most severe scenario for climate impacts of a nuclear winter (a 61% reduction in food production), diverted food exports could still provide 1.5 times the current dietary energy intakes for the whole NZ population (or 13,298 kJ [3178 kcal] per person per day). In the much smaller regional nuclear war scenario (5 Tg of stratospheric soot, 8% reduction in food production), diverted food exports could still provide 3.6 times the current dietary energy intakes of the population.

## Discussion

### Main findings and interpretation

This preliminary analysis suggests that NZ’s current food production for export is theoretically able to provide an excess of dietary energy for the whole population, even after a severe nuclear winter that reduced food production across-the-board by 61%. As such, this baseline excess food production capacity would also be a resilience factor after other sunlight-reducing planetary catastrophes such as large volcanic eruptions and asteroid/comet impacts. Furthermore, given the breadth of food production in NZ, there are no survival-critical food products that would be missing from the national diet after such catastrophes.

Despite the preliminary estimates in this study considering climate effects of nuclear winter on agricultural production, the agriculture sector is very much interconnected with other key systems in modern technological society. Therefore, there is a need for further research on all interlinked domains considered by Zeihan,^45^ in addition to international trade and societal functioning, as detailed in Table 6.

**Table 6.** Domains (other than the agriculture) of relevance to food security in a post-nuclear war/nuclear winter setting

| Domain<br>(largely adapted from Zeihan<sup>45</sup>) | Potential details relevant to post-nuclear war food security requiring further research |
| --- | --- |
| Transport | Food is transported internally in NZ by rail, truck and van. Except for the North Island Main Trunk railway and suburban rail systems (which are electrified), these transport modes are highly dependent on imported liquid fossil fuels. The exploratory work in NZ on electric-milk tankers<sup>46</sup> and hydrogen-powered trucks<sup>47</sup> is still at a very early stage. In a post-war setting, some existing electric cars and vans could be re-purposed for food delivery, but possibly food production would need to be intensified closer to cities and towns. |
| Finance | An operational financial system in NZ could be threatened by nuclear war, with a risk of collapse.<sup>24</sup> Unemployed and retired people would need money to buy food from farmers, food processors and food distributors. Therefore, central and local governments might need to have backup food rationing systems and systems for prioritising food supplies to essential workers and those at greatest need. Fortunately, the NZ Government obtained some valuable experience with rapidly providing mass welfare support during the Covid-19 pandemic – but the scale of a post-nuclear war situation would probably be vastly greater and longer lasting.<br>Appropriate financing may also be needed to assist the agricultural sector in diverting crops suitable for human food that are currently used for animal feed and other purposes. For example, an estimated 78% of cereals produced in NZ are used for livestock feed (ie, much of the wheat, maize and oats).<sup>48</sup> Furthermore, some of the remaining 22% of cereals could be used more efficiently to feed humans eg, if more of the barley crop was used for making flour rather than being used to make beer and spirits. Fodder beet (a type of beetroot) and forage brassicas (eg, canola, radish, turnip, swede, and kale) could also be converted from stock feed to human food. Alternatively, this forage cropping land could be used for other food crops. |
| Energy | In addition to energy for transport (see above), energy is needed to run agricultural machinery. Some of this is electrical energy (eg, for irrigation systems and milking sheds), but much of the machinery uses imported diesel fuel. Substitutes for this diesel are fairly minimal with low levels of biofuel production (made from whey, tallow, and used cooking oil<sup>49</sup>). Liquid biofuels may need to be prioritised for the most efficient food crops (for dietary energy per hectare) such as grains and potatoes – and perhaps sectors such as fisheries would have to miss out.<br>A further risk to electrical grids (and telecommunications) in NZ is that an electromagnetic pulse (EMP) from nuclear attacks on facilities Australia might have spill-over impacts on NZ. The probability of such attacks on Australia in a nuclear war are highly uncertain, as is also the impact of EMP itself (it is very dependent on the altitude of the burst).<sup>50</sup> |
| Industrial materials | Many inputs into agriculture are imported, including much of the fertiliser, pesticides, animal health pharmaceuticals, and spare parts for agricultural machinery. NZ is a major seed exporter for some food crops (eg, radishes, carrots and beets<sup>51</sup>) but is also reliant on imports. |
| Manufacturing | Although NZ does make some of its own nitrogen fertiliser, it is dependent on imports for phosphate-based fertiliser.<sup>52</sup> The capacity to manufacture replacements for imported materials (as per those in the row immediately above) is uncertain. |
| Trade (international) | If post-war trade was able to be re-established with regional neighbours such as Australia and Indonesia, then these countries could provide some liquid fuels and machinery parts used for food production in NZ. If so, NZ could strive for some ongoing food exports to these countries so that imports could be paid for. Trade with Pacific island nations could include the export of coconut oil, sugar and dried fish to NZ, with coconut oil also being a potential diesel substitute. |
| Societal functioning | Given all the potential impacts referred to in this table, especially if there is an end to international trade and risk of financial collapse, NZ society could be extremely shocked after a Northern Hemisphere nuclear war. Factors such as the competence of central and local government and the quality of communication by societal leaders would also probably be very important. This is a complex area to research (with relevant NZ writing being fairly out-of-date<sup>53</sup>), but new work could build on studies of the other domains in this Table, and studies of how NZ and other societies has responded to other shocks (including the Covid-19 pandemic).<br>In terms of refugees from other countries, it seems plausible that the difficulties with access to shipping in a post-war environment could substantially limit the arrival of such refugees. Nevertheless, a full analysis of food supply issues under a range of plausible circumstances might give an indication of how many refugees NZ could readily absorb. It is also possible that with imported fuel shortages refugees could help with providing additional manual labour for food production. |

### Study strengths and limitations

A strength of this preliminary study is that it is the first one to take such an in-depth bottom-up look at food security in NZ after a nuclear winter. That is, other analyses have taken a higher level approach to dietary energy availability in the business-as-usual case^54^ (ie, 9569 kcal/per person/day in 2013 for NZ), and in the post-nuclear-winter case.^26,28^ Nevertheless, our work is still preliminary given we have not explored all the important other sectors that agriculture, food processing and food delivery are interlinked with (as per Table 6). Other notable limitations of our analysis include the following:

- It does not consider the size of NZ’s non-export food economy – due to its vastly greater complexity. This domain includes the food produced for the domestic market, household level production (eg, vegetable gardening and on lifestyle blocks), and citizen harvesting food from the environment (eg, fishing, hunting and shellfish gathering). Nevertheless, one estimate for 2018 was that the country produced 7768 kilotonnes of food, of which 2216 kilotonnes (28.5%) was for the domestic market. However, this previous estimate did not consider stocks from previous years and production waste (ie, losses during transportation and storage).^48^ Also, our analysis only considered major food export domains by ignoring average annual food exports of under 10,000 tonnes (ie, excluding such food exports as: beans, berry fruits, brassicas, capsicums, carrots, citrus, eggs, honey, pears, squash, and tomatoes).^48^
- The nuclear winter impacts involved various simplifying assumptions. For example, the estimates from Xia et al^28^ were for selected crops and marine fish and we extrapolated from these to across-the-board reductions in food productivity. Although the model by Xia et al considered impacts on “surface air temperature, precipitation and downward direct and diffuse solar radiation”, it did not consider potential damage to agriculture from increased ultra-violet light after a nuclear war.^55^ There may also be complex nuclear winter effects on sea-ice and oceans that impact on fisheries for very long periods.^29^
- The nutrition analysis was only for dietary energy and so further work could consider the scope for achieving nutritionally-balanced diets. Nevertheless, protein availability would not seem to be a major concern, given the current extent of exported dairy products, meat, and marine products. NZ is also self-sufficient in egg production, and produces grains and legumes that are protein sources (eg, beans for the export market).^48^

### Potential implications for further research and resilience building

Given the uncertainties, further work into food security after nuclear winter and other sunlight-reducing scenarios is desirable, and could focus particularly on the issues detailed in Table 6. More specific research domains within the agricultural sector are detailed in Table A1 in the Appendix.Some of these also have the potential co-benefit of strengthening the resilience of NZ’s food systems, following Tendall et al’s definition of food systems resilience, “including social, economic and biophysical processes operating at many scales”.^56^

## Conclusions

This analysis suggests that NZ could have excess food production capacity, even after a severe nuclear winter scenario. But substantial further research is needed to clarify agricultural impacts and the role of nuclear war impacts on the interlinked domains of energy, transport, manufacturing, finance, industrial materials, trade and societal functioning.

## Data Availability

All key data are within the paper. The Excel file with specific food energy calculations is available from the authors on request.

## Data Availability

All key data are within the paper. The Microsoft Excel file with specific food energy calculations is available from the authors on request.

## Acknowledgements

Nil.

## Appendix

**Table A1.** Potential research areas for enhancing food security resiliency in NZ after a nuclear winter and other sunlight reducing catastrophes

| Domain | Additional details |
| --- | --- |
| **Food production** |  |
| Enhanced soil protection | Research could consider the most cost-effective approach to enhance soil protection – especially of crop land near cities. Enhanced protection of soils has previously been argued for NZ on general food security grounds.<sup>48</sup> Options include increasing tree planting on erosion-prone land and better control of introduced mammalian pests (eg, deer, pigs and goats). The protection of high-quality soils from urban development may also contribute to greater resilience and a national policy statement for the protection of the 15% of the country’s land considered to be “highly productive” was produced in 2022.<sup>57</sup> Reducing soil erosion would also help protect other food sources: shellfish and other kaimoana (seafood) in estuaries, and mahinga kai (eg, eels) in rivers and streams. |
| Increased electrification of agriculture and food systems | Research could explore how to make the agricultural sector less reliant on imported fossil fuels eg, via increasing numbers of electric vehicles and greater electrification of farm machinery and food processing machinery. Using hydro-generated electricity for hydrogen fuel production (eg, for trucks and trains) in NZ,<sup>58</sup> could be given further attention. Although not as energy efficient as electric and diesel vehicles, hydrogen could still provide an additional level of resilience. |
| Greater agricultural self-sufficiency | Research could study ways to support the agricultural sector to become less reliant on imported fertilisers and imported feeds (eg, palm kernel expeller). |
| Optimising land-use | Research could examine how to support “best fit” land-use change so that farming systems are optimally suited to local soils, geography, and climate. This could enhance food security and resiliency to climatic shocks.<sup>59</sup> |
| Increased urban food production | Research could explore how to increase urban food production by households (eg, fruit, vegetables, and poultry), in community gardens, and by local Māori agribusiness that can supply food directly to iwi members.<sup>60</sup> The Government-funded think tank Te Puna Whakaaranui has argued that “empowering communities to produce and distribute food is a low cost, culturally inclusive approach to food security”.<sup>61</sup> This think tank cites local examples such as the Hauora Kai Co-Op<sup>62</sup> and WELLfed.<sup>63</sup> Priority crops to promote (from a nuclear winter perspective), could be ones that are high in dietary energy per land area required eg, potatoes. Potatoes also have moderate tolerance to frost.<sup>64</sup> There are also frost-tolerant vegetables such as onions, turnips and beets (especially sugar beet as frost hardiness is correlated with high sugar content).<sup>64</sup> |
| Seed banks | Research could examine the value of seed banks for seeds for frost-tolerant and frost-resistant crops and seeds that NZ currently imports. |
| **Differing types of food and food use** |  |
| Plant-based diets | Research could explore how to best promote a faster shift to more plant-based diets as these are typically more sustainable, lower cost and healthier as per various NZ studies.<sup>65-67</sup> |
| Enhanced marine food supply | Research could consider the expansion of marine food reserves through better management of coastal fisheries and expansion of the number and size of marine reserves. It has been estimated that “...effective prewar management that rebuilds fish biomass could ensure a short-term catch buffer large enough to replace ~43 ± 35% of today's global animal protein production”.<sup>27</sup> Increased consumption of discarded bycatch and marine offal, could also increase food supplies for people in a post-war setting. Research is already underway of the expansion of commercial seaweed production in NZ<sup>68</sup> and this food source could potentially be scaled up in many coastal areas. |
| Food waste reduction | Research could further examine ways to reduce food waste at all levels in the food system, given the NZ data on the substantial size of this problem.<sup>36</sup> Reducing household level food waste now could have immediate cost-saving benefits for citizens and reduce greenhouse gas footprints. |

